# Establishing the causal role of intimate partner violence and abuse on depressive symptoms in young adults: a population-based cohort study

**DOI:** 10.1101/2021.06.07.21258467

**Authors:** Annie Herbert, Jon Heron, Maria Barnes, Christine Barter, Gene Feder, Khadija Meghrawi, Eszter Szilassy, Abigail Fraser, Laura D Howe

## Abstract

**Background:** Previous studies have shown an association between experience of intimate partner violence and abuse (IPVA) and depression. Whether this is a causal relationship or explained by prior vulnerability that influences the risk of both depression and IPVA is not known.

**Methods and Findings:** We analysed data from the Avon Longitudinal Study of Parents and Children prospective cohort (N=1,764 women, 1,028 men). To assess the causal association between IPVA at 18-21 years old and logged depressive symptom scores at age 23, we used: i) multivariable linear regression, ii) inverse probability of treatment weighting (IPTW), and iii) difference-in-difference (DID) analysis, which compared the mean change in logged depressive symptom scores between ages 16 and 23 between those who experienced IPVA and those who did not.

Women who experienced IPVA had on average 26% higher depressive symptom score after adjustment for measured confounders (ratio of geometric means 1.26, 95% CI 1.13 to 1.40). In men, the difference was 5% (ratio of geometric means 1.05, 95% CI 0.92 to 1.21). Results from IPTW analysis were similar. In the DID analysis, there was no evidence that being exposed to IPVA affected the change in depressive symptom scores over time compared to being in the non-exposed group for either women (difference-in-differences: 1%, -12% to 16%) or men (−1%, -19% to 20%).

**Conclusions:** Multivariable linear regression and IPTW suggested an association between IPVA and higher depressive symptom score in women but not men, but DID analysis indicated a null effect in both women and men. This suggests the causal origins of higher depressive symptoms in this young adult population are likely to reflect prior vulnerability that leads to both higher depressive symptoms and increased risk of IPVA exposure.

**Key messages:**

- Women and, to a lesser degree, men who experience IPVA between the ages of 18-21 years old tend to have higher levels of depressive symptoms at age 23 years than people who do not.
- In women, the association between IPVA and higher depressive symptoms remained after adjustment for measured confounders or use of inverse probability of treatment weighting, whereas in men, the association attenuated to the null after adjustment for measured confounders or use of inverse probability of treatment weighting.
- However, the change in depressive symptoms between ages 16 and 23 years was the same in people who experience IPVA and people who did not, for both women and men. This suggests that the higher levels of depressive symptoms in people who experience IPVA is likely to be driven by prior vulnerability that affects the risk of both IPVA exposure and depressive symptoms, highlighting that IPVA is one of a series of challenges being navigated by psychologically vulnerable young adults.

## Introduction

Intimate partner violence and abuse (IPVA) is experienced by approximately 30% of women and 24% of men between 18 and 21 years of age in the UK.(1) IPVA may have lasting consequences for mental health, yet there is a lack of long-term studies on the effect of IPVA on depressive symptoms. Although longitudinal studies investigating the relationship between IPVA and mental health problems, particularly depression, consistently report bidirectional associations, they usually do not have strong study designs and robust analytical approaches for assessing causality.(2) Confounding may influence these findings if background factors such as adversity in childhood influence both the risks of experiencing IPVA and depressive symptoms. Moreover, the majority of previous studies combine findings from young and middle-aged adults.

Some longitudinal evidence is available from younger adolescent samples,(3) suggesting an increase in depressive symptoms following IPVA, at five years following IPVA, after adjusting for baseline depression.(4, 5) The one study that additionally adjusted for other potentially confounding factors (race, age, socioeconomic status, childhood maltreatment, and pubertal status) reported small increases in mean Centre for Epidemiology Studies Depression scores at five years following psychological or physical IPVA, were reported.(4, 5) However, the number and range of factors adjusted for in this analysis may not fully capture the large range of factors that can influence both IPVA and depression. Therefore, concerns remain about the extent to which associations between IPVA and depressive symptoms in young adults are causal. This is important to establish because it has implications for interventions. If associations are not causal, while prevention of IPVA and support to survivors is still essential, we also need effective interventions in earlier stages of the life course to prevent adult mental health problems.

Here, we use data from a UK general population-based birth cohort to investigate the longitudinal relationship between IPVA at age 18-21 and depressive symptoms at age 23, using three techniques - multivariable linear regression, inverse probability of treatment weighting, and a difference-in-difference model.

## Methods

### Data and participants

We analysed data from the Avon Longitudinal Study of Parents and Children (ALSPAC) cohort. Around ∼14,500 pregnant women residing in Avon, UK, with expected delivery dates in April 1991-December 1992 (approximately three-quarters of the eligible population) were recruited. When the oldest children were approximately 7 years of age, an attempt was made to bolster the initial sample with eligible cases who had failed to join the study originally, resulting in an additional 913 children being enrolled. Information has been regularly collected since enrolment until present. Study data were collected and managed using REDCap electronic data capture tools hosted at University of Bristol.(6) More information on both the mothers and their offspring is available in published cohort profiles.(7-9). The study website contains details of all the data that is available through a fully searchable data dictionary and variable search tool.(10)

Within questionnaires offered in both online and paper format, 3,280 ALSPAC participants answered questions on IPVA at age 21 (interquartile range for age of response: 21 to 22). These questions ask about IPVA both prior to turning 18 and between age 18 and 21. In order to make causal inferences, it was important to establish the timing of exposure to IPVA such that we could account for confounding; therefore, we excluded participants exposed to IPVA prior to turning 18 (n=487) leaving a final analysis sample of 2,793 participants.

### Exposure: IPVA

At age 21, ALSPAC participants were asked about IPVA, capturing psychological, physical, and sexual IPVA. For example, how often an intimate partner had ‘Told you who you could see and where you could go and/or regularly checked what you were doing and where you were (by phone or text)?’, to which one could respond ‘never’, ‘once’, ‘a few times’, or ‘often’, and whether this occurred prior to turning 18, after turning 18, or at both time-points. These questions been previously developed based on previous UK and European questionnaires and the PROVIDE questionnaire,(11, 12) and are described in full in a report of their psychometric properties.(13) As in previous work, we considered any response of at least ‘once’, to any of the eight questions as exposure to IPVA, because the header of the questionnaire was *‘Intimate Partner Violence’*, likely raising the threshold of severity for reporting certain behaviours, and because for participants who answered at least ‘once’ to any of the questions, negative impact was reported by 75–99%.(1)

We also included different IPVA types (psychological, physical, sexual) in analyses. The questions used to distinguish different types have been described previously.(13) Combinations of types were grouped based on both the sample prevalence of different combinations, and existing literature finding variation in impact and mental health between such combinations.(14, 15) The categories of IPVA types analysed were: no victimisation, psychological victimisation only, physical victimisation (whether with or without psychological victimisation, no sexual), and any sexual victimisation.

### Outcome: Depressive symptom scores

Depressive symptoms were captured at age 23 via Moods and Feelings (MFQ) questionnaire scores. This questionnaire asks 13 questions about depressive symptoms in the past two weeks, with response options ‘not true’, ‘somewhat true’ or ‘true’ (scoring 0, 1, and 2, respectively).

### Other variables

Covariates were included in models either to estimate adjusted coefficients, or to create propensity scores (described later under ‘Inverse probability of treatment weighting’).

These covariates were known risk factors for IPVA and depressive symptoms (i.e. could confound the relationship between the two) based on previous empirical literature, or, as recommended by the propensity score methods literature,(16, 17) variables that were known risk factors for, as a minimum, the outcome (i.e. depressive symptoms).(18, 19) These were all coded as binary variables: socioeconomic status at birth [Index Multiple Deprivation quintiles 4-5 vs. 1-3], ethnicity [White vs. Person of Colour], sexual minority at age 15 [100% heterosexual vs. other], depression at age 16, anxiety at age 17, extreme parental monitoring at age 15, anti-social behaviour at age 14, smoking at age 16 [at least weekly], cannabis use at age 16 [at least weekly], illicit (non-cannabis) drug use at age 16 [any past month], hazardous alcohol use at age 18, adverse childhood experiences at ages 0-16 [ten different variables: emotional abuse, physical abuse, sexual abuse, emotional neglect, bullying, witnessing domestic violence, parental mental health problem, parental substance abuse, parental criminal conviction, and parental separation],(18, 20-22) low self-esteem at age 17 (Bachman Self Esteem Scale score <=29),(23) overweight at age 17 (body mass index >=25), sleep problems at age 17 (Clinical Interview Schedule-Revised sleep subsection score >=2),(24) and parents’ education level (at least one parent with at least O level qualifications).(13)We also included a measure of depressive symptoms prior to exposure (MFQ score at age 16, as having prior depressive symptoms is a risk factor for IPVA and later depression). More detail on how the above variables were derived using ALSPAC data has been published previously.(1, 25)

### Statistical analysis methods

We carried out all analyses separately for women and men, given that some outcomes of IPVA have been shown to differ by sex, e.g. reported impact of IPVA (1). As per disclosure rules for use of ALSPAC data, we do not report any numbers (or related percentages) less than 5. R scripts used in these analyses are available at: https://github.com/pachucasunrise/IPVA_depression.

We employed the following three methods to test the causality of the effect of IPVA on depression.

#### 1. Multi-variable linear regression

We fitted a model where depressive symptom score at age 23 was the dependent variable, and IPVA at age 18-21 was the independent variable. An unadjusted model was compared with two adjusted models: ‘Adjusted Model A’ included as covariates variables that had been adjusted for within existing literature (4, 5): MFQ score at age 16, ethnicity, socioeconomic status and four ACEs representing child maltreatment (emotional abuse, physical abuse, sexual abuse, emotional neglect). Adjusted model B included all covariates listed in the ‘Other variables’ section.

#### 2. Inverse probability of treatment Weighting (IPTW)

A linear regression was fitted where the outcome was depression score at age 23, and (IPVA at 18-21 vs. none) was included as an independent variable. Participants were weighted using stabilised weights calculated as a function of the propensity to experience IPVA (estimated from a separate logistic regression model that included all relevant measured covariates as independent variables) and the global probability of IPVA.(26) Further information on IPTW and the methods used are in **Supplementary Box S1**.

#### 3. Difference-in-Differences (DID)

Here, we fitted a linear regression model where each individual contributed two data-points on the ‘outcome’ (depression score), one at 16 years, before the time-period for exposure, and one after at age 23.(27-29) The assumption is that the difference between groups (IPVA at age 18-21 vs. none) in depressive symptom scores at baseline (prior to exposure) represents all confounding, both measured and unmeasured. The difference in the change in depression scores between 16-23 years between those exposed to IPVA and those not exposed is then assumed to reflect the causal effect of IPVA on change in depression score, under a set of assumptions. Further information on DID and the methods used are described in **Supplementary Box S2**.

We repeated the above three methods for categories of victimisation types. For IPTW, we used multinomial logistic regression to estimate propensity scores for different categories. For DID analysis, we fitted three difference models for each of the categories of victimisation types vs. no victimisation.

We use the natural log of depressive symptom scores due to its non-normal distribution. The exponentiated regression coefficient for a logged outcome represents the ratio of geometric means. Using logged depressive symptom scores, residual histograms were adequately Normally distributed and p-values from the Kolmogorov-Smirnoff test for the fully adjusted model were 0.90 and 0.72 for women and men respectively.

To facilitate comparison with the existing literature, we present the odds ratio for the association of IPVA with a dichotomised measure of depression (defined as depressive symptom score above 12).

### Missing data

Participants were included in analyses if they had data on depressive symptom scores at ages 16 and 23, IPVA exposure, and ethnicity. For other covariates, we imputed any missing values using multiple imputation via chained equations, separately for women and men. We imputed 50 datasets using the MICE package in R,(30) with 10 iterations. Regression models were fitted in each of these imputed datasets and coefficients and standard errors pooled using Rubin’s rules.(31) Where p-values were reported, this was the median p-value between the 50 imputed datasets.

For IPTW, within each imputed dataset propensity scores and stabilised weights were estimated and regressions inverse-weighted, before pooling resulting coefficients across the imputed datasets.(32)

### Sensitivity analyses

We repeated the main analysis within completely observed data, and again using the non-transformed variable of depressive symptom scores.

The main analysis was restricted to those who reported no IPVA victimisation before age 18. People exposed to IPVA both before age 18 and at age 18-21 may differ in other ways to those who were only exposed at age 18-21. We repeated analyses without this restriction (N = 3,280 vs. 2,793). However, it must be noted that we can now no longer assume that the variables adjusted for or the difference in mean depression score at age 16 occurred prior to the IPVA exposure. Therefore, we cannot make causal inference from those results.

We also repeated analyses restricting to the 90% of individuals who indicated that they had had an intimate relationship/encounter by age 21 (N = 2,421). This was to more robustly meet the positivity assumption of causality (i.e., that all individuals in the sample can be exposed to IPVA).

To assess the common trends assumption for DiD analysis, we checked that differences in mean logged depressive symptom scores were constant between the ages 13 and 16 for people who did and did not report IPVA victimisation at 18-21 years.

## Results

### Cohort characteristics

Of 1,764 women and 1,028 men in the study cohort, 482 (27%) and 210 (20%) reported that they had been victimised between age 18 and 21, respectively; characteristics of participants according to whether or not they experienced IPVA victimisation are presented in **Supplementary Table S1**.

### Relationship between victimisation and subsequent depression scores

The geometric mean depressive symptom scores at age 23 was 5.2 for both women and men in the non-victimised group, and 7.0 and 6.2 in women and men who experienced IPVA victimisation. In women, IPVA was associated with a doubling in the odds of depressive symptoms above the threshold for defining depression (OR 2.10, 95% CI 1.57 to 2.81). In men, IPVA was associated with a 36% increase in the odds of depression (OR 1.36, 95% CI 0.91 to 2.04) (**Table 1**).

**Table 1.**
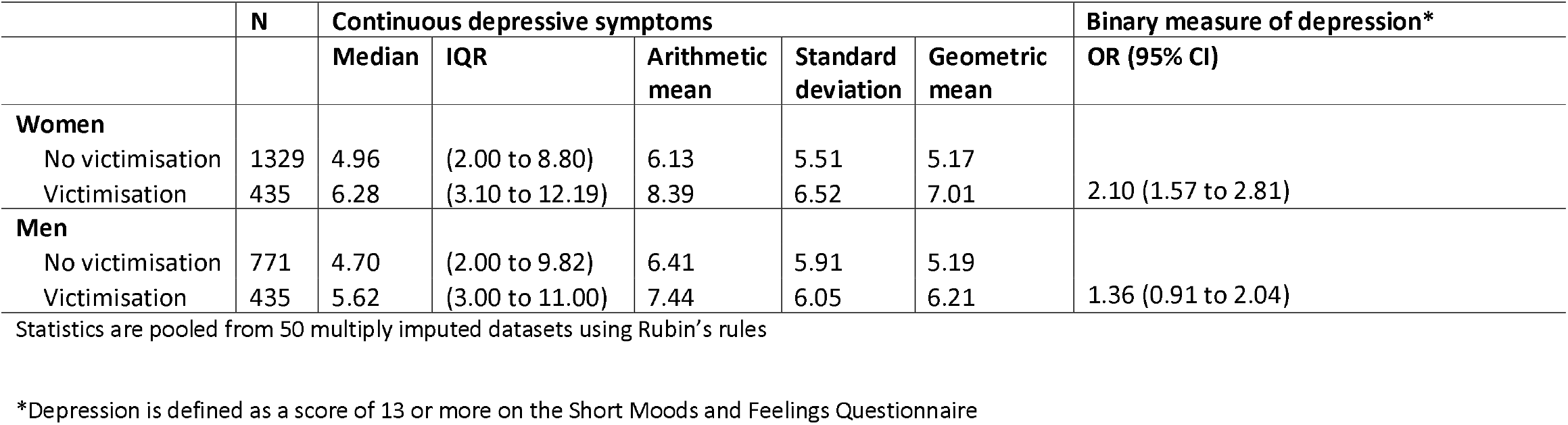
Statistics on depressive symptom scores and a binary depression measure at age 23, stratified by sex and reporting of victimisation at age 18-21.

#### Multivariable linear regression

Depressive symptoms at age 23 were 34% higher in women and 20% higher in men who reported being victimised at age 18-21 compared to those who had not reported being victimised (ratio of geometric means in women: 1.34, 95% CI 1.21 to 1.49; men: 1.20, 1.04 to 1.38) (Table 2, crude models). The estimated difference in depressive symptom scores between victimised and non-victimised groups was reduced after adjusting for logged depressive symptom scores at age 16, ethnicity, socioeconomic status, and certain ACEs; the association still remained in women (26%; ratio of geometric means 1.26, 95% CI 1.13 to 1.40), but was attenuated to the null in men (6%; ratio of geometric means 1.06, 95% CI 0.92 to 1.21) (**Table 2**, Adjusted Model A); these results remained relatively unchanged after adjusting for the larger set of covariates in Adjusted Model B.

**Table 2.** Association between IPVA victimisation at age 18-21 and logged depressive symptom score at age 23 using linear regression and IPTW. Analysis on multiply imputed data, N=1764 women and 1028 men.

|  | **Women** |  | **Men** |  |
| --- | --- | --- | --- | --- |
| **Model** | **% change** | **Ratio of geometric means (95% CI)** | **% change** | **Ratio of geometric means (95% CI)** |
| 1 (Crude) | 36 | 1.36 (1.23, 1.51) | 20 | 1.20 (1.04, 1.37) |
| 2 (Adjusted A*) | 26 | 1.26 (1.13, 1.40) | 6 | 1.06 (0.92, 1.21) |
| 3 (Adjusted B**) | 26 | 1.26 (1.13, 1.40) | 5 | 1.05 (0.92, 1.21) |
| 4 (IPTW) | 20 | 1.20 (1.01, 1.43) | 5 | 1.05 (0.84, 1.32) |

Depressive symptom score was logged, and the coefficients from linear regression models were exponentiated to ratios of geometric means, and subsequently converted to percent changes in the geometric mean of depressive symptom score; % change = $[\exp(\text{coefficient})-1]*100$ . Statistics are pooled from 50 multiply imputed datasets using Rubin's rules, as described in Methods. IPTW = Inverse probability of treatment weighting. \*Adjusted for logged depressive symptom score at age 16, ethnicity, socioeconomic status, and dummy variables for childhood emotional abuse, physical abuse, sexual abuse, and emotional neglect.
\*\*As in Adjusted A\* and additionally adjusted for sexual minority status, anxiety, extreme parental monitoring, anti-social behaviour, smoking, cannabis use, illicit (non-cannabis) drug use, hazardous alcohol use, bullying, witnessing domestic violence, parental mental health problem, parental substance abuse, parental criminal conviction, and parental separation.

#### IPTW

When regression estimates were inverse probability of treatment weighted, associations between IPVA and depressive symptoms were in line with the multivariable adjusted results (women: 20%; ratio of geometric means 1.20, 95% CI 1.01 to 1.43; men: 5%, ratio of geometric means 1.05, 95% CI 0.92 to 1.21).

#### DID

Depressive symptoms prior to IPVA victimisation (age 16) were 29% higher in women who went on to experience IPVA victimisation between 18-21 years (ratio of geometric means 1.29, 95% CI 1.17 to 1.42), and 34% higher in men who went on to experience IPVA victimisation between 18-21 years (ratio of geometric means 1.34, 95% CI 1.16 to 1.55) (**Table 3; Figure 1**). Depressive symptom scores increased over time in men, but les so in women. There was little evidence of a difference in differences (i.e., that being in the victimised group affected the change in geometric mean depressive symptom scores over time compared to being in the non-victimised group; the interaction between victimisation and time), for either women (2%, ratio of geometric means 1.02, 95% CI 0.89 to 1.18, p-value=0.76) or men (−5%, ratio of geometric means 0.95, 95% CI 0.78 to 1.18, p-value=0.67).

**Table 3.** Difference-in-difference analysis for the relationship between IPVA victimisation at age 18-21 and logged depressive symptom scores at ages 16 and 23 (models fitted in imputed datasets, N = 1764 women and 1028 men).

| Term | Women |  |  | Men |  |  |
| --- | --- | --- | --- | --- | --- | --- |
|  | % change<sup>Ⓜ</sup> | Ratio of geometric means (95% CI) | p-value | % change<sup>Ⓜ</sup> | Ratio of geometric means (95% CI) | p-value |
| Victimisation<sup>‡</sup> | 29 | 1.29 (1.17, 1.42) | <0.001 | 34 | 1.34 (1.16, 1.55) | <0.001 |
| Time<sup>‡‡</sup> | 5 | 1.05 (0.98, 1.13) | 0.18 | 34 | 1.34 (1.22, 1.47) | <0.001 |
| Victimisation*Time<sup>‡‡‡</sup> | 2 | 1.02 (0.89, 1.18) | 0.76 | -5 | 0.95 (0.77, 1.18) | 0.665 |

CI = Confidence interval
<sup>Ⓜ</sup>ln geometric mean. % change calculated from the estimated coefficient for victimisation in each model, as % change = [exp(coefficient)-1]\*100.
<sup>‡</sup> The difference in depressive symptoms at baseline (age 16 years) comparing people exposed to IPVA with people not exposed to IPVA
<sup>‡‡</sup> The change in depressive symptoms over time, in people not exposed to IPVA
<sup>‡‡‡</sup> The difference in difference, i.e. the difference in the change in depressive symptoms over time, comparing people exposed to IPVA with people not exposed to IPVA.

**Figure 1.**
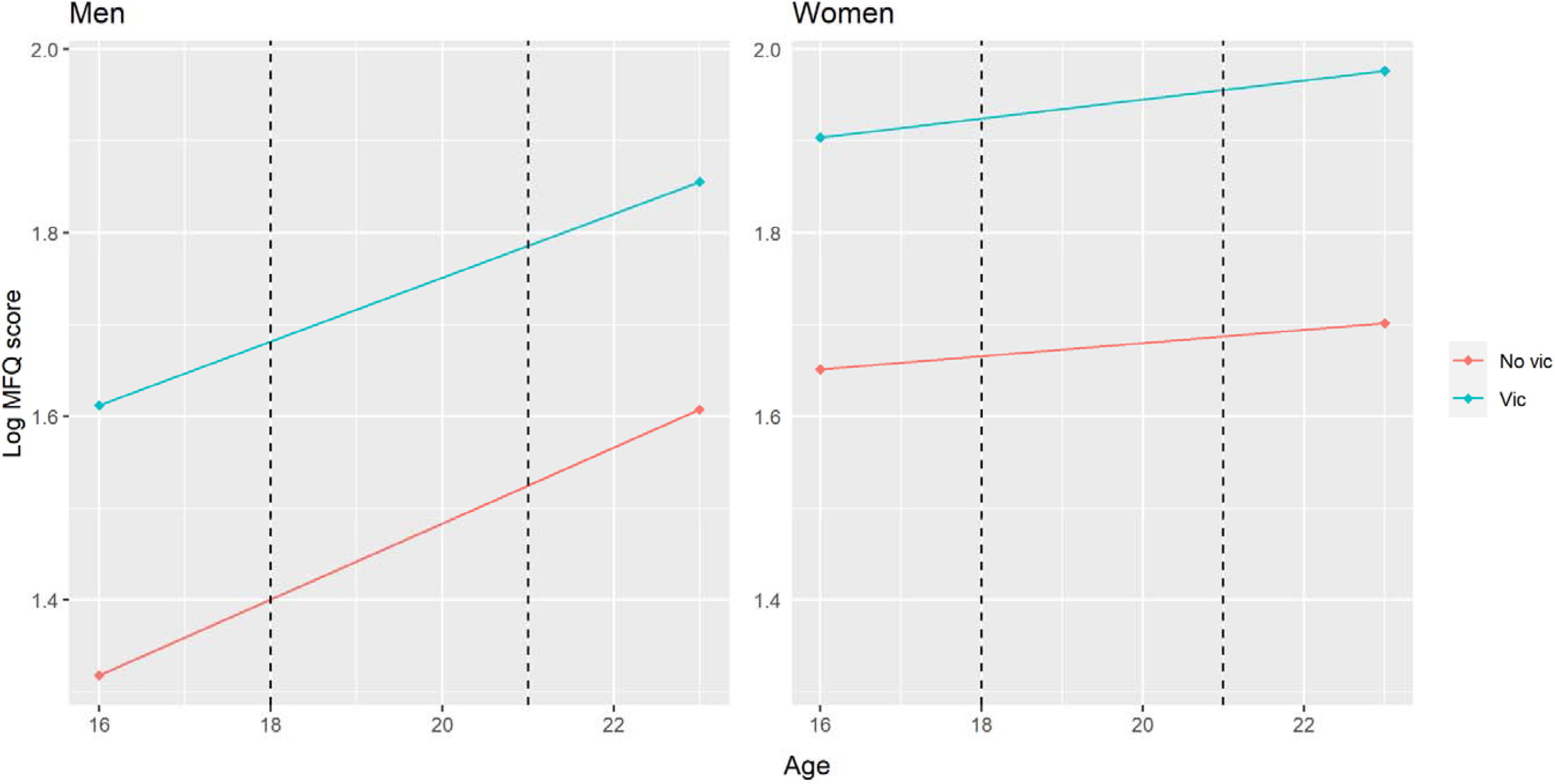
Estimates from difference-in-difference analysis for the relationship between IPVA victimisation at age 18-21 and logged depressive symptom scores at ages 16 and 23. Dashed lines represent ages 18-21 (when the exposure IPVA was reported to occur).

### Relationship between different categories of victimisation types and subsequent depression scores

Of 1,764 women and 1,028 men in the study cohort, 209 women (12%) and 123 men (12%) reported being psychologically victimised only at ages 18-21, 111 (6%) and 50 (5%) reported being physically victimised, with or without experiencing psychological victimisation but without sexual victimisation, and 162 (9%) and 37 (4%) reported being sexually victimised.

#### Multivariable linear regression

The geometric mean was 22% and 5% higher for women and men who were psychologically victimised only, 37% and 20% higher for women and men physically victimised (whether psychologically victimised or not, but not sexually victimised), and 55% and 47% higher for women and men who were sexually victimised (**Table 4**). These associations roughly halved after adjustment. Most associations were imprecisely estimated due to the small number of people experiencing each subtype of victimisation.

**Table 4.** Regression estimates for the relationship between different categories of IPVA types at age 18-21 and logged depressive symptom score at age 23 (models fitted in imputed data, N=1764 women and 1028 men).

| Model | Subtype of IPVA | Women |  | Men |  |
| --- | --- | --- | --- | --- | --- |
|  |  | % change | Ratio of geometric means (95% CI) | % change | Ratio of geometric means (95% CI) |
| 1 (Crude) | Psychological only | 22 | 1.22 (1.06, 1.41) | 5 | 1.05 (0.87, 1.28) |
|  | Physical (with or without psych, no sex) | 37 | 1.37 (1.13, 1.66) | 20 | 1.20 (0.92, 1.57) |
|  | Any sexual | 55 | 1.55 (1.29, 1.87) | 47 | 1.47 (1.17, 1.86) |
| 2 (Adjusted A*) | Psych only | 25 | 1.25 (1.08, 1.44) | -4 | 0.96 (0.79, 1.16) |
|  | Phys (with or without psych, no sex) | 31 | 1.31 (1.08, 1.58) | 12 | 1.12 (0.87, 1.43) |
|  | Any sexual | 22 | 1.22 (1.01, 1.48) | 20 | 1.20 (0.96, 1.50) |
| 3 (Adjusted B**) | Psych only | 28 | 1.28 (1.11, 1.48) | -4 | 0.96 (0.79, 1.16) |
|  | Phys (with or without psych, no sex) | 29 | 1.29 (1.06, 1.56) | 10 | 1.10 (0.86, 1.41) |
|  | Any sexual | 18 | 1.18 (0.98, 1.43) | 20 | 1.20 (0.96, 1.50) |
| 4 (IPTW) | Psych only | 19 | 1.19 (1.02, 1.39) | 0 | 1.00 (0.81, 1.24) |
|  | Phys (with or without psych, no sex) | 11 | 1.11 (0.90, 1.38) | -3 | 0.97 (0.72, 1.30) |
|  | Any sexual | 1 | 1.01 (0.83, 1.23) | 43 | 1.43 (1.13, 1.81) |

Any sexual = Any sexual victimisation (with or without psychological or physical victimisation); CI = Confidence interval; IPTW = Inverse probability of treatment weighting;
Phys = Physical victimisation (with or without psychological, no sexual); Psych only = Psychological victimisation only
Statistics are pooled from 50 multiply imputed datasets using Rubin's rules, as described in Methods. % change calculated from the estimated coefficient for victimisation category in each model, as % change = $[\exp(\text{coefficient}) - 1] * 100$ .
\*Adjusted for logged depressive symptom score at age 16, ethnicity, socioeconomic status, and dummy variables for childhood emotional abuse, physical abuse, sexual abuse, and emotional neglect.
\*\*As in Adjusted A\* and additionally adjusted for sexual minority status, anxiety, extreme parental monitoring, anti-social behaviour, smoking, cannabis use, illicit (non-cannabis) drug use, hazardous alcohol use, bullying, witnessing domestic violence, parental mental health problem, parental substance abuse, parental criminal conviction, and parental separation.

#### IPTW

When regression estimates were inverse probability of treatment weighted, most estimates attenuated (**Table 4**). One exception was the estimate for sexual victimisation in men (which increased from a 20% higher geometric mean depressive symptom scores in Adjusted Model B to 43% in the IPTW model, 95% CI for the ratio of geometric means 1.13 to 1.81).

#### DID

There was no evidence that any victimisation subtype affected the change in depressive symptoms over time, for either women or men (Table 5).

**Table 5.**
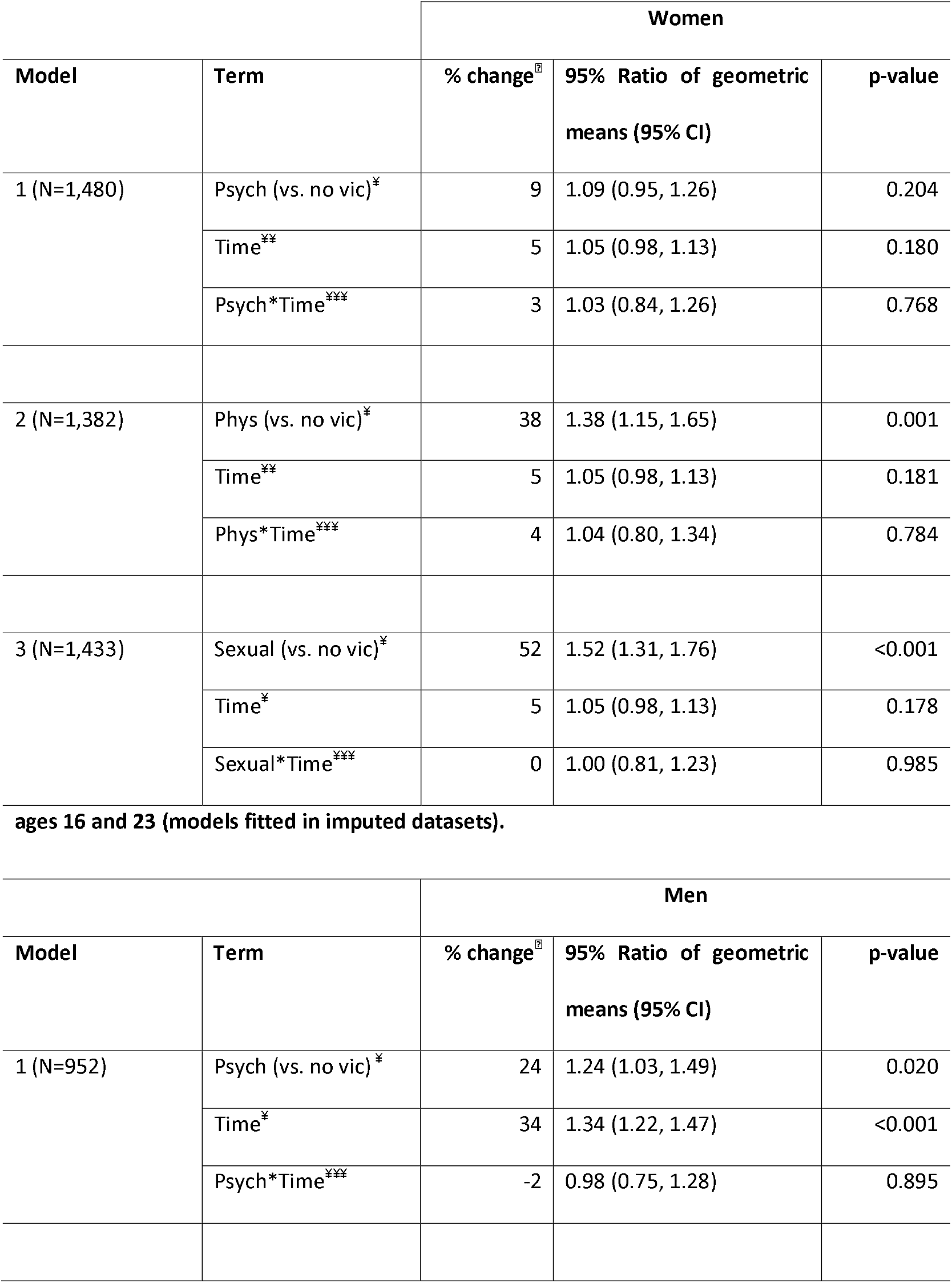
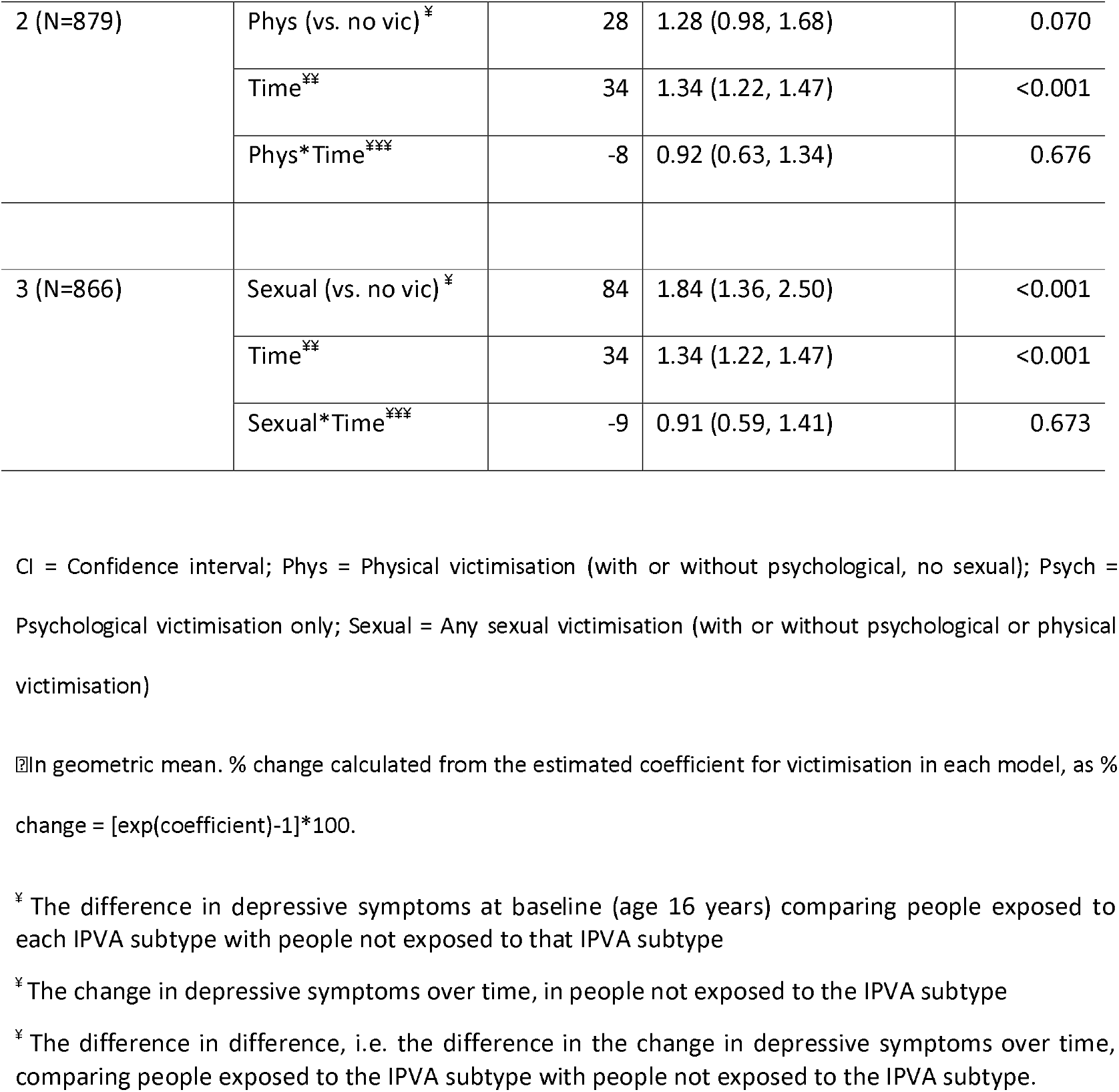
Difference-in-difference analyses for the relationships between different categories of victimisation types at age 18-21 (vs. no victimisation) and logged depressive symptom scores at.

|  |  | **Women** |  |  |
| --- | --- | --- | --- | --- |
| **Model** | **Term** | **% change<sup>®</sup>** | **95% Ratio of geometric means (95% CI)** | **p-value** |
| 1 (N=1,480) | Psych (vs. no vic)<sup>‡</sup> | 9 | 1.09 (0.95, 1.26) | 0.204 |
|  | Time<sup>‡‡</sup> | 5 | 1.05 (0.98, 1.13) | 0.180 |
|  | Psych*Time<sup>‡‡‡</sup> | 3 | 1.03 (0.84, 1.26) | 0.768 |
| 2 (N=1,382) | Phys (vs. no vic)<sup>‡</sup> | 38 | 1.38 (1.15, 1.65) | 0.001 |
|  | Time<sup>‡‡</sup> | 5 | 1.05 (0.98, 1.13) | 0.181 |
|  | Phys*Time<sup>‡‡‡</sup> | 4 | 1.04 (0.80, 1.34) | 0.784 |
| 3 (N=1,433) | Sexual (vs. no vic)<sup>‡</sup> | 52 | 1.52 (1.31, 1.76) | <0.001 |
|  | Time<sup>‡</sup> | 5 | 1.05 (0.98, 1.13) | 0.178 |
|  | Sexual*Time<sup>‡‡‡</sup> | 0 | 1.00 (0.81, 1.23) | 0.985 |

**ages 16 and 23 (models fitted in imputed datasets).**
|  |  | **Men** |  |  |
| --- | --- | --- | --- | --- |
| **Model** | **Term** | **% change<sup>®</sup>** | **95% Ratio of geometric means (95% CI)** | **p-value** |
| 1 (N=952) | Psych (vs. no vic)<sup>‡</sup> | 24 | 1.24 (1.03, 1.49) | 0.020 |
|  | Time<sup>‡</sup> | 34 | 1.34 (1.22, 1.47) | <0.001 |
|  | Psych*Time<sup>‡‡‡</sup> | -2 | 0.98 (0.75, 1.28) | 0.895 |
| 2 (N=879) | Phys (vs. no vic)<sup>‡</sup> | 28 | 1.28 (0.98, 1.68) | 0.070 |
|  | Time<sup>‡‡</sup> | 34 | 1.34 (1.22, 1.47) | <0.001 |
|  | Phys*Time<sup>‡‡‡</sup> | -8 | 0.92 (0.63, 1.34) | 0.676 |
| 3 (N=866) | Sexual (vs. no vic)<sup>‡</sup> | 84 | 1.84 (1.36, 2.50) | <0.001 |
|  | Time<sup>‡‡</sup> | 34 | 1.34 (1.22, 1.47) | <0.001 |
|  | Sexual*Time<sup>‡‡‡</sup> | -9 | 0.91 (0.59, 1.41) | 0.673 |

CI = Confidence interval; Phys = Physical victimisation (with or without psychological, no sexual); Psych = Psychological victimisation only; Sexual = Any sexual victimisation (with or without psychological or physical victimisation)
<sup>‡‡</sup>In geometric mean. % change calculated from the estimated coefficient for victimisation in each model, as % change = [exp(coefficient)-1]\*100.
<sup>‡</sup> The difference in depressive symptoms at baseline (age 16 years) comparing people exposed to each IPVA subtype with people not exposed to that IPVA subtype
<sup>‡‡</sup> The change in depressive symptoms over time, in people not exposed to the IPVA subtype
<sup>‡‡‡</sup> The difference in difference, i.e. the difference in the change in depressive symptoms over time, comparing people exposed to the IPVA subtype with people not exposed to the IPVA subtype.

### Sensitivity analyses

Analyses using the crude depressive symptom score at age 23 (not logged) followed a similar pattern to when the outcome was logged (**Supplementary Tables S2 and S3**).

When complete case data were analysed, or when we did not restrict the cohort according to whether reporting victimisation by age 17, or when we analysed data only on participants who reported an intimate relationship by age 17; patterns in estimates from Adjusted Model B, the IPTW model and DiD analyses were similar compared to the main analyses (**Supplementary Table S4 and S5**). There were exceptions; the IPTW models when we included participants who had reported victimisation before age 18 indicated an increase of 43% (ratio of geometric means 1.43, 95% CI 1.29 to 1.58) among women and 26% (ratio of geometric means 1.26, 95% CI 1.10 to 1.44) among men. However, the estimate from DiD analysis including participants who had reported victimisation before age 18 was similar to the main analysis.

There was no evidence to suggest a violation of the common trends assumption; p values for interaction between IPVA victimisation status and changes in depressive symptom score between 13-16 years were 0.33 for women and 0.59 for men (**Supplementary Box S2**).

## Discussion

Average depression scores at age 23 were 1-2 points higher for women and men who reported being victimised at age 18-21 compared to those who did not amongst both women and men in a UK population-based birth cohort. When controlling for measured confounding using either multivariable linear regression or IPTW, analyses indicated a small effect of IPVA victimisation on depressive symptom scores in women but not men. In contrast, DID analyses, which can account for unmeasured time-fixed confounding, suggested no causal effect in either women or men. On balance, although women and men victimised at age 18-21 were more vulnerable to depression at age 23, the associations observed in multivariable regression and IPTW analyses are likely affected by mismeasured and/or unmeasured confounders, and it is likely that the causal component of this relationship is either small or null, and mainly driven by prior vulnerability that increased propensity to be exposed to IPVA. Sensitivity analyses suggested that the assumptions of the DiD analysis were met. However, even if the null effect of the DID analyses was driven by violated assumptions, the maximum possible causal effect suggested by regression analyses would be around 0.9 points on the MFQ scale, a small effect, given a standard deviation of 5 to 6 points. This effect is much less than an assumed minimum important clinical difference of 5 points on the MFQ scale.(33)

### Strengths and limitations

We analysed data on a large population-based cohort, using validated measures of IPVA and depression. A strength of this cohort is the availability of a range of different risk factors for IPVA and depression, and of depression itself, at multiple time-points. This allowed us to carry out both IPTW and DID, which has not been possible in previous datasets. Internal checks found no evidence of violation of the common trends assumption for DiD analysis, but if this assumption was violated, we would anticipate it would likely bias the effect away from the null.

Depression scores were captured at age 23, 2-5 years following measured exposure to IPVA (depending on whether the participant was closer to age 18 or 21 at the time of exposure). It is possible that some participants did not develop depressive symptoms related to their IPVA exposure until after age 23. Similarly, as the IPVA may have occurred several years prior to measurement of depressive symptoms, some participants may have had recovering mental health by this time. A review of the published literature on trajectories of psychopathology following potentially traumatic events such as child maltreatment, cancer diagnosis, heart attack, or bereavement, indicated that an average of 65.7% of participants across studies experienced a ‘resilience’ trajectory, 20.8% experienced recovery, 10.6% experienced chronic symptoms, and 8.9% experienced delayed onset of symptoms.(34)

There is a possibility that selection bias may have distorted effect estimates. Around one-third of individuals still in the cohort at 21 years old responded to the age 21 questionnaire. Given previous methodological work on the ALSPAC cohort indicating that those experiencing poorer socioeconomic and health outcomes will be less likely to participate,(35) we could expect those with the worst depressive states and IPVA experiences would be less likely to participate. Indeed, individuals who did not respond to the age 21 questionnaire tended to have lower depressive symptom scores at age 16 than those who had responded; it was not possible to check this phenomenon for IPVA as it was not measured prior to age 21. If this were the case, and those with the worst IPVA experiences were less likely to participate, this would attenuate the effect of IPVA on depression, and so the ‘true’ general young adult population effect may in fact be larger. However, the extent of any bias is uncertain given that the cohort still includes individuals with high depressive symptom scores (10% had scores of 10-12) or IPVA experiences,(15, 36) in our data.

### Comparison with literature

A systematic review and meta-analysis of longitudinal studies identified 16 studies with 36,163 participants(2). Most of those studies were not restricted to young adults. All but one study identified a positive association between IPVA and incident depressive symptoms in women, with a pooled odds ratio of 1.97 (95% CI 1.56-2.48). There was also evidence of an association between IPV and depression in men, but only two studies included data for men so meta-analysis was not performed. Our main analysis used a continuous measure of depressive symptoms, which is not directly comparable to the binary measures used in this meta-analysis, but when we dichotomised our measure of depressive symptoms the odds ratio for the relationship between IPVA and depression was similar to the pooled result from the meta-analysis for both women and men, supporting the notion that our findings are not influenced by selection bias. In line with our conclusion about prior vulnerability influencing the association between IPVA and depressive symptoms, the meta-analysis also found evidence of an association between depressive symptoms and incident IPV (pooled odds ratio from four studies 1.93, 95% CI 1.51-2.48).

Our findings are comparable with one of the only longitudinal studies we could identify that studied depression following IPVA in young people.(5) In the National Longitudinal Survey of Adolescent Health (NLSAH) which surveyed a younger US population (aged 12-18) about psychological and physical IPVA, depression was captured using a different measure but with similarly worded questions and was on a similar scale,(37, 38), and with similar follow-up. The authors found a difference of up to 0.9 points (depending on IPVA sub-type, 95% CIs ranging from 0.01 to 1.76) after adjustment for confounders equivalent to those we adjusted for in Adjusted Model A.(5) In this study, the associations of IPVA with depression were similar for women and men, although gender differences were seen for other outcomes.

### Implications

Our findings indicate the likely mental health detriment in the years following IPVA exposure in a UK young adult population. The higher levels of depressive symptoms experienced by both women and men who experience IPVA victimisation are likely to be associated with other adverse health and social outcomes.(39-43) However, our findings suggest that IPVA may not be a direct cause of the higher burden of depressive symptoms in people who experience IPVA, thus highlighting that IPVA is one of a series of challenges being managed by psychologically vulnerable young people. In addition to supporting young adults who experience IPVA, with strategies for recognising and ending unhealthy relationships, these young adults would likely benefit from support in other areas of their life and life history.

Our results also have implications for further research. We adjusted models in two stages, and found that estimates only marginally altered when we adjusted for variables beyond those typically adjusted for in previous literature: prior depression scores, ethnicity, socioeconomic status, childhood emotional abuse, physical abuse, sexual abuse, and emotional neglect.(4, 5) However, DID analyses accounts for unmeasured confounders, and here even evidence of small effects of IPVA on depression scores disappeared. We recommend that a DID analysis is applied to other longitudinal IPVA and mental health data to investigate whether our findings can be replicated in other populations, as there may be other risk factors for IPVA and depression or effect modifiers that are not being measured in relevant cohorts. Future exploration of such factors, including through qualitative interviews to capture processes not often or easily measured in quantitative studies, could strengthen our understanding of risk factors for experiencing IPVA.(3, 44)

When we examined the relationship between IPVA and depression for different victimisation categories, confidence intervals for the effect of sexual victimisation were wide. However, the point estimate was large (36%; 95% CI: -14% to 113%) for men but not for women (9%, 95% CI: -6% to 26%). It is possible that the mental health impacts across different categories of IPVA are more similar for women than for men, as women are far more likely than men to suffer frequent ‘multi-victimisation’ or ‘intimate terrorism’.(15, 45) Further exploration of the mental health impacts of sexual abuse in young men and women using larger samples, such as existing multi-country studies in adolescents,(46) and older adult women,(47) is warranted.

## Conclusion

In a UK general population sample, young people experiencing intimate partner violence are likely to have more depressive symptoms than those not victimised. Addressing these symptoms needs to take into account possible exposure to IPVA, and the mental health needs of young survivors of IPVA need to be addressed in the context of IPVA services. However, the causal origins of this increased susceptibility to depression are principally explained by prior vulnerability that increases both depressive symptoms and the risk of IPVA exposure.

### Consent and ethical approval

Written informed consent was obtained from the parents of participating children after receiving a full explanation of the study. Children were invited to give assent where appropriate. Study members have the right to withdraw their consent for elements of the study or from the study entirely at any time. Full details of the ALSPAC consent procedures are available on the study website (http://www.bristol.ac.uk/alspac/researchers/research-ethics/). The questions on IPVA were approved by the ALSPAC Ethics and Law Committee (ref: E201210).

## Data Availability

ALSPAC data access is through a system of managed open access. The steps below highlight how to apply for access to ALSPAC data, including access to the Stata/R scripts used for analyses reported in this Research Article.

http://www.bristol.ac.uk/alspac/researchers

## Data availability statement

1. Please read the ALSPAC access policy, available from http://www.bristol.ac.uk/alspac/researchers/access/, which describes the process of accessing the data and samples in detail, and outlines the costs associated with doing so.
2. You may also find it useful to browse our fully searchable <u>research proposals database</u>, which lists all research projects that have been approved since April 2011.
3. Please <u>submit your research proposal</u> for consideration by the ALSPAC Executive Committee. You will receive a response within 10 working days to advise you whether your proposal has been approved.

If you have any questions about accessing data, please.

The ALSPAC data management plan describes in detail the policy regarding data sharing, which is through a system of managed open access.

## Acknowledgements

We are extremely grateful to all the families who took part in the ALSPAC study, the midwives for their help in recruiting them, and the whole ALSPAC team, which includes interviewers, computer and laboratory technicians, clerical workers, research scientists, volunteers, managers, receptionists and nurses. We are grateful to Drs Ana Luiza Goncalvez Soares and Gemma Clayton who advised on IPTW and DID analyses, respectively. We are grateful to members of our project advisory group for valued discussion on the findings, the interpretation, and implications, including Amanda Flanagan (Respect), John Devaney (Edinburgh University), and Jayne Whittlestone (Next Link).

## Author contributions

AH, JH, AF and LDH devised the study. AH carried out statistical analyses and wrote the first draft of the manuscript. All authors contributed to study design, interpretation of results, and critical revisions of the manuscript.

