## Supplementary materials for "Establishing the causal role of intimate partner violence and abuse on depressive symptoms in young adults: a population-based cohort study"

Annie Herbert^1,2^, Jon Heron^1,2^, Maria Barnes^1,3^, Christine Barter^4^, Gene Feder^1,3^, Eszter Szilassy^1,3^, Abigail Fraser^1,2,5^, Laura D Howe^1,2^

1 – Department of Population Health Sciences, University of Bristol, Bristol, UK

2 – MRC Integrative Epidemiology Unit at the University of Bristol, Bristol, UK

3 – Centre for Academic Primary Care, University of Bristol, Bristol, UK

4 – University of Central Lancashire, Preston, UK

5 – NIHR Biomedical Research Centre, University of Bristol, UK

**Supplementary Box S1: Inverse Probability Treatment Weighting (IPTW)**

For Inverse Probability Treatment Weighting (IPTW), a model is fitted to determine an individual’s ‘propensity’ to be exposed to IPVA based on their set of covariate values. One issue with estimating weights is that they can become unstable if the probability of exposure to IPVA is either very close to 0 or 1. Therefore, we used ‘stabilised weights’.(1) The stabilised weight for an individual $i$ in group $j$ is:

$$s_{i}= \frac{p}{\pi_{i}} if {IPVA}_{i}=1$$

$$= \frac{p}{(1-\pi_{i})} if {IPVA}_{i}=0$$

where $i$=1,…N, $\pi_{i}$ = probability of exposure to IPVA given individual $i$’s set of covariate values, $p$ = probability of exposure to IPVA. The logistic regression model used to estimate $\pi_{i}$ includes IPVA as the dependent variable and for independent variables, covariates considered to be associated with either exposure and outcome, or at least the outcome.(2) **Figure S1** shows the distribution of propensity scores between exposure groups for men and women, within the first imputed dataset. The analysis model is fitted, where individuals are weighted according to $s_{i}$.

In IPTW we assume conditional exchangeability (exchangeability conditional on the inverse propensity scores) as all relevant confounders have been included in the model used to estimate propensity scores, such that the distribution of covariates between groups are similar, i.e., after weighting the non-IPVA and IPVA group become comparable. This can be checked by comparing standardised differences of covariates before and after weighting (a standardised difference of <0.05 indicating adequate covariate balance), which are shown in **Figure S2**. An advantage of propensity score methods such as IPTW is that the model for estimating propensity scores was separate and so the potential for over-fitting with a large number of covariates to consider would not affect the final analysis model (which is the case for multivariable linear regression). Often estimates from IPTW are similar to those for multivariable regression where the two methods have considered the same covariates, but confidence intervals will differ depending on how the outcome is distributed among covariates.(3, 4) Among the possible propensity score methods (including matching and stratification), we chose IPTW as it can be used to estimate the Average Treatment Effect in the whole population, it does not discard any observations, and it performs well when combined with imputation methods.(2) Finally, there is a large advantage of IPTW over other propensity score methods in that, standardised differences in covariates before and after weighting could be checked to explicitly assess whether covariates were balanced conditional on re-weighting (indicating some level of exchangeability).

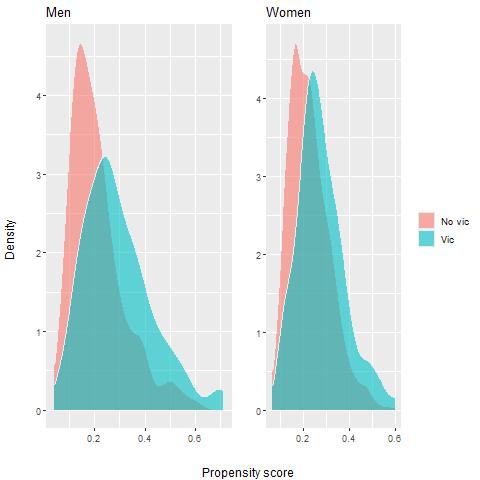

**Figure S1. Propensity score distributions between those exposed to IPVA at age18-21 and those not, within the first imputation.**

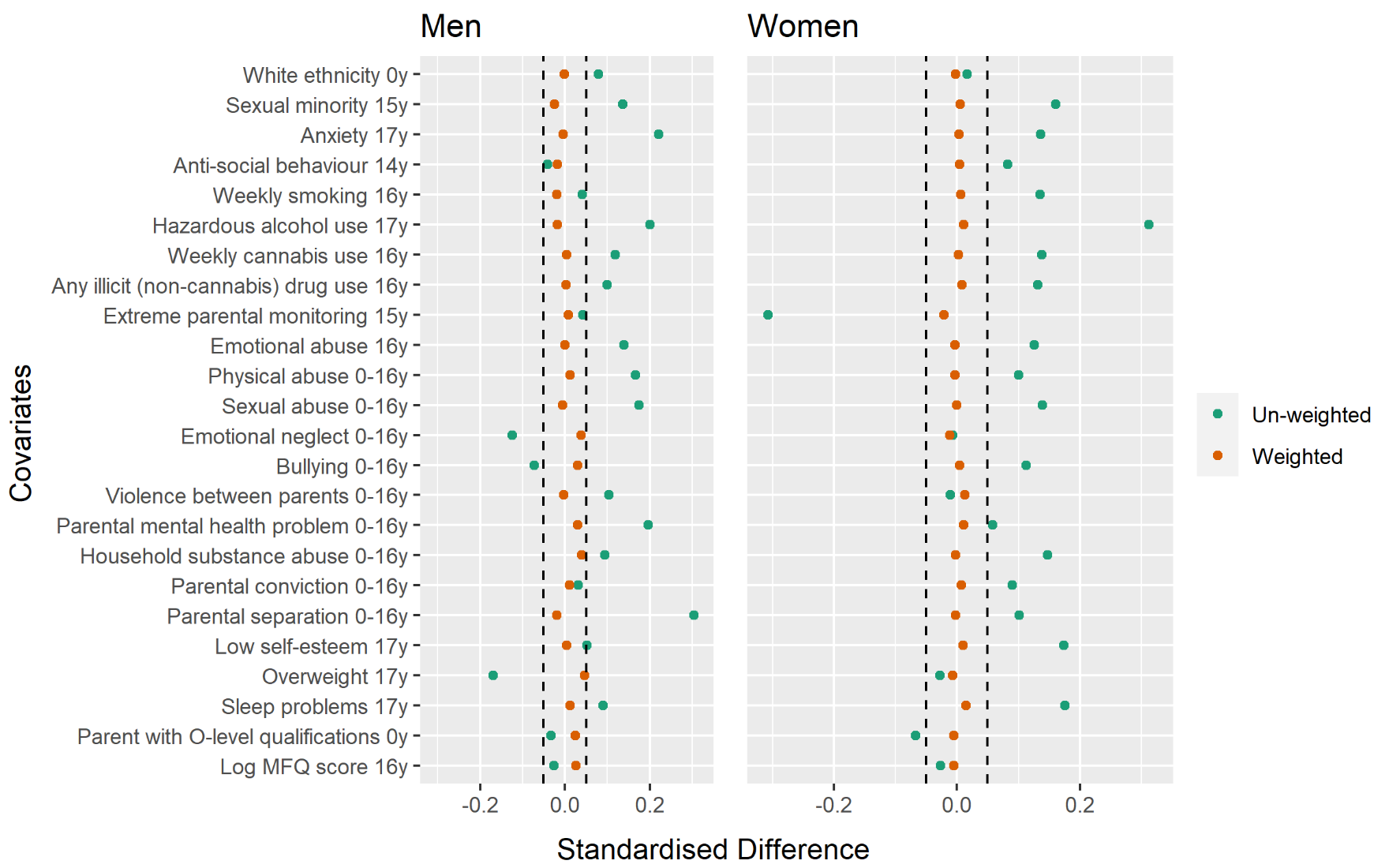

**Figure S2. Standardised differences in proportions* between those exposed to IPVA at age18-21 and those not, before and after weighting by stabilised inverse propensity scores**

*Except for log depressive symptom score where the standardised mean difference is presented.

**Supplementary Box S2: Difference-in-difference (DID) analysis**

Differences in mean scores at age 16 are assumed to represent differences caused by all possible confounders, including unmeasured ones (e.g. attitudes to reporting experiences such as mental health problems or IPVA experiences).(5, 6) Therefore, any further difference in mean scores after the exposure (i.e. in mean scores at age 23) between groups, must represent a causal effect. The following model was fitted:

$$y_{ij}=\beta_{0}+\beta_{1}T_{ij}+\beta_{2}I_{i}+\beta_{3}I_{i}T_{ij}+e_{ijt}$$

where $y =$ logged depressive symptom score; $i = 1,\ldots.n$, the index per individual; $j = 0,1$ the index for repeated observations per individual; $T = 0,1$ is a dummy variable indicating whether the observation occurred before (=0) or after (=1) age 18-21; $I = 0,1$ is a dummy variable indicating whether or not exposed to IPVA at age 18-21; $\beta_{0}$ = the mean logged depressive symptom score at age 16 for those not exposed to IPVA at age 18-21; $\beta_{1}$ = the difference in mean logged depressive symptom scores between age 16 and age 23 for those not exposed to IPVA at age 18-21; $\beta_{2}$ = the difference in mean logged depressive symptom scores at age 16 between those not exposed to IPVA at age 18-21, and those exposed; $\beta_{3}$ = the difference-in-differences of mean logged depressive symptom scores at age 16 and age 23 between those not exposed to IPVA at age 18-21, and those exposed. That is, the ‘difference-in differences’, interpreted as the causal effect of IPVA exposure on depression is the coefficient for the interaction between IPVA grouping and time, $\beta_{3}$.

Individuals contributed two data-points each in the model, and so robust standard errors were used to account for the correlation between data-point pairs.

In DID analysis we assume conditional exchangeability (exchangeability conditional on the difference in logged depressive symptom scores at baseline) by assuming that the initial difference in logged depressive symptom scores between the non-IPVA and IPVA groups prior to the exposure time-point (i.e. at age 16) represents the effect of all measured and unmeasured time-invariant confounders on the outcome, and that there is no time-varying confounding between groups (e.g. socio-economic status does not have a smaller/larger effect on depression scores at age 23 compared to at age 16), also known as the ‘common trends assumption’.(7) For this to be possible, we also have to assume ; no variation in measurement error between groups or over time (e.g. any difference between depressive symptom score and actual depressive symptomatology will not be affected by whether age 16 or 23, or whether in the non-IPVA or IPVA group).(6) To assess the common trends assumption, we checked that differences in mean logged depressive symptom scores were constant between the ages 13 and 16. There was no evidence to suggest that there was an interaction between IPVA grouping and changes in logged depressive symptom scores between ages 13 and 16 (p-value for men = 0.59; women = 0.33).

**Table S1: Characteristics of the cohort before age 18, by those victimised at 18-21 and not, and sex**

|  |  | **Males (n=1039)** | | | | **Females (n=1753)** | | | |
| --- | --- | --- | --- | --- | --- | --- | --- | --- | --- |
|  |  | **No victimisation** | | **Victimisation** | | **No victimisation** | | **Victimisation** | |
| **Variable (age in years)** | **Level** | **n** | **(%)** | **n** | **(%)** | **n** | **(%)** | **n** | **(%)** |
| N |  | 829 | (100) | 210 | (100) | 1271 | (100) | 482 | (100) |
| **Demographics** |  |  |  |  |  |  |  |  |  |
| Index of Multiple Deprivation (0) | Quintiles 1-3 (less deprived) | 470 | (56.7) | 116 | (55.2) | 707 | (55.6) | 236 | (49) |
|  | Quintiles 4-5 (more deprived) | 143 | (17.2) | 37 | (17.6) | 207 | (16.3) | 85 | (17.6) |
|  | Missing | 216 | (26.1) | 57 | (27.1) | 357 | (28.1) | 161 | (33.4) |
| Ethnicity (0) | White | 793 | (95.7) | 199 | (94.8) | 1205 | (94.8) | 452 | (93.8) |
|  | Non-white | 26 | (3.1) | 10 | (4.8) | 55 | (4.3) | 22 | (4.6) |
|  | Missing | 10 | (1.2) | 1 | (0.5) | 11 | (0.9) | 8 | (1.7) |
| Sexual minority (15) | No | 580 | (70) | 128 | (61) | 803 | (63.2) | 270 | (56) |
|  | Yes | 56 | (6.8) | 20 | (9.5) | 90 | (7.1) | 48 | (10) |
|  | Missing | 193 | (23.3) | 62 | (29.5) | 378 | (29.7) | 164 | (34) |
| **Mental and physical health** |  |  |  |  |  |  |  |  |  |
| Depression score (16)* | *Median (IQR)* | *3* | *(1 to 5)* | *4* | *(2 to 7)* | *4* | *(2 to 9)* | *6* | *(3 to 10)* |
|  | Missing | 193 | (23.3) | 58 | (27.6) | 240 | (18.9) | 123 | (25.5) |
| Anxiety (17) | No | 544 | (65.6) | 129 | (61.4) | 698 | (54.9) | 236 | (49) |
|  | Yes | 17 | (2.1) | 10 | (4.8) | 60 | (4.7) | 29 | (6) |
|  | Missing | 268 | (32.3) | 71 | (33.8) | 513 | (40.4) | 217 | (45) |
| Low self-esteem score (17)* | No | 339 | (40.9) | 79 | (37.6) | 420 | (33) | 124 | (25.7) |
|  | Yes | 250 | (30.2) | 67 | (31.9) | 491 | (38.6) | 203 | (42.1) |
|  | Missing | 240 | (29) | 64 | (30.5) | 360 | (28.3) | 155 | (32.2) |
| Sleep problems (17)* | No | 486 | (58.6) | 115 | (54.8) | 675 | (53.1) | 226 | (46.9) |
|  | Yes | 111 | (13.4) | 33 | (15.7) | 190 | (14.9) | 97 | (20.1) |
|  | Missing | 232 | (28) | 62 | (29.5) | 406 | (31.9) | 159 | (33) |
| Overweight (17)* | No | 469 | (56.6) | 126 | (60) | 667 | (52.5) | 253 | (52.5) |
|  | Yes | 120 | (14.5) | 21 | (10) | 195 | (15.3) | 70 | (14.5) |
|  | Missing | 240 | (29) | 63 | (30) | 409 | (32.2) | 159 | (33) |
| **Risk-taking behaviours** |  |  |  |  |  |  |  |  |  |
| Anti-social behaviour (14) | No | 660 | (79.6) | 154 | (73.3) | 1017 | (80) | 348 | (72.2) |
|  | Yes | 56 | (6.8) | 11 | (5.2) | 78 | (6.1) | 35 | (7.3) |
|  | Missing | 113 | (13.6) | 45 | (21.4) | 176 | (13.8) | 99 | (20.5) |
| Smoking (16) | No | 636 | (76.7) | 152 | (72.4) | 991 | (78) | 334 | (69.3) |
|  | Yes | 33 | (4) | 9 | (4.3) | 95 | (7.5) | 47 | (9.8) |
|  | Missing | 160 | (19.3) | 49 | (23.3) | 185 | (14.6) | 101 | (21) |
| Hazardous alcohol use (17) | No | 367 | (44.3) | 81 | (38.6) | 563 | (44.3) | 167 | (34.6) |
|  | Yes | 189 | (22.8) | 60 | (28.6) | 239 | (18.8) | 139 | (28.8) |
|  | Missing | 273 | (32.9) | 69 | (32.9) | 469 | (36.9) | 176 | (36.5) |
| Cannabis (16) | No | 653 | (78.8) | 154 | (73.3) | 1076 | (84.7) | 377 | (78.2) |
|  | Yes | 16 | (1.9) | 7 | (3.3) | 8 | (0.6) | 8 | (1.7) |
|  | Missing | 160 | (19.3) | 49 | (23.3) | 187 | (14.7) | 97 | (20.1) |
| Illicit (non-cannabis) drug use (16) | No | 650 | (78.4) | 153 | (72.9) | 1043 | (82.1) | 361 | (74.9) |
|  | Yes | 18 | (2.2) | 6 | (2.9) | 38 | (3) | 23 | (4.8) |
|  | Missing | 161 | (19.4) | 51 | (24.3) | 190 | (14.9) | 98 | (20.3) |
| **Parental factors** |  |  |  |  |  |  |  |  |  |
| At least one parent O-level qualification (birth) | No | 94 | (11.3) | 26 | (12.4) | 180 | (14.2) | 80 | (16.6) |
|  | Yes | 735 | (88.7) | 184 | (87.6) | 1091 | (85.8) | 402 | (83.4) |
| Extreme parental monitoring (15) | No | 403 | (48.6) | 92 | (43.8) | 473 | (37.2) | 222 | (46.1) |
|  | Yes | 267 | (32.2) | 66 | (31.4) | 457 | (36) | 116 | (24.1) |
|  | Missing | 159 | (19.2) | 52 | (24.8) | 341 | (26.8) | 144 | (29.9) |
| **Adverse Childhood Experiences** |  |  |  |  |  |  |  |  |  |
| Emotional abuse (0-11) | No | 588 | (70.9) | 130 | (61.9) | 856 | (67.3) | 279 | (57.9) |
|  | Yes | 129 | (15.6) | 40 | (19) | 185 | (14.6) | 81 | (16.8) |
|  | Missing | 112 | (13.5) | 40 | (19) | 230 | (18.1) | 122 | (25.3) |
| Physical abuse (0-11) | No | 588 | (70.9) | 131 | (62.4) | 873 | (68.7) | 288 | (59.8) |
|  | Yes | 122 | (14.7) | 41 | (19.5) | 176 | (13.8) | 74 | (15.4) |
|  | Missing | 119 | (14.4) | 38 | (18.1) | 222 | (17.5) | 120 | (24.9) |
| Sexual abuse (0-16) | No | 766 | (92.4) | 182 | (86.7) | 1097 | (86.3) | 385 | (79.9) |
|  | Yes | 6 | (0.7) | 6 | (2.9) | 41 | (3.2) | 26 | (5.4) |
|  | Missing | 57 | (6.9) | 22 | (10.5) | 133 | (10.5) | 71 | (14.7) |
| Emotional neglect (0-16) | No | 550 | (66.3) | 142 | (67.6) | 830 | (65.3) | 310 | (64.3) |
|  | Yes | 157 | (18.9) | 30 | (14.3) | 173 | (13.6) | 62 | (12.9) |
|  | Missing | 122 | (14.7) | 38 | (18.1) | 268 | (21.1) | 110 | (22.8) |
| Bullying (8-16) | No | 516 | (62.2) | 139 | (66.2) | 858 | (67.5) | 309 | (64.1) |
|  | Yes | 201 | (24.2) | 46 | (21.9) | 193 | (15.2) | 92 | (19.1) |
|  | Missing | 112 | (13.5) | 25 | (11.9) | 220 | (17.3) | 81 | (16.8) |
| Witnessed domestic violence (0-12) | No | 570 | (68.8) | 133 | (63.3) | 782 | (61.5) | 284 | (58.9) |
|  | Yes | 99 | (11.9) | 30 | (14.3) | 171 | (13.5) | 58 | (12) |
|  | Missing | 160 | (19.3) | 47 | (22.4) | 318 | (25) | 140 | (29) |
| Parental mental illness or suicide attempt (0-16) | No | 455 | (54.9) | 92 | (43.8) | 636 | (50) | 211 | (43.8) |
|  | Yes | 272 | (32.8) | 83 | (39.5) | 414 | (32.6) | 154 | (32) |
|  | Missing | 102 | (12.3) | 35 | (16.7) | 221 | (17.4) | 117 | (24.3) |
| Parental substance abuse (0-11) | No | 661 | (79.7) | 152 | (72.4) | 957 | (75.3) | 316 | (65.6) |
|  | Yes | 54 | (6.5) | 17 | (8.1) | 68 | (5.4) | 37 | (7.7) |
|  | Missing | 114 | (13.8) | 41 | (19.5) | 246 | (19.4) | 129 | (26.8) |
| Parental criminal conviction (0-12) | No | 683 | (82.4) | 161 | (76.7) | 988 | (77.7) | 339 | (70.3) |
|  | Yes | 40 | (4.8) | 11 | (5.2) | 54 | (4.2) | 26 | (5.4) |
|  | Missing | 106 | (12.8) | 38 | (18.1) | 229 | (18) | 117 | (24.3) |
| Parental separation (0-16) | No | 556 | (67.1) | 111 | (52.9) | 773 | (60.8) | 251 | (52.1) |
|  | Yes | 128 | (15.4) | 50 | (23.8) | 215 | (16.9) | 87 | (18) |
|  | Missing | 145 | (17.5) | 49 | (23.3) | 283 | (22.3) | 144 | (29.9) |

Data presented as ns and column %s.

*Depression score = Moods & Feelings Questionnaire score reported within postal/online questionnaire at age 16 (questions are listed in Table 8 of Angold *et al* (1995) (8, 9); Low self esteem score = Bachman-Rosenberge-Extended self esteem score <=29 reported via online/postal questionnaire at age 17 (questions are listed in Table 7-2 of Bachman *et al* (1970) (10, 11); Sleep problems = Sleep symptom score >= 2 reported in clinic at age 17 in the sleep sub-section of the Clinical Interview Schedule-Revised (CIS-R) (12, 13); Overweight = BMI >= 25 measured in clinic at age 17. How all other variables were derived is described in detail in Herbert *et al* (2020), Extended data, Table B.(14)

**Table S2. Comparison of estimated coefficients from different regressions investigating the relationship between IPVA victimisation at age 18-21 and depressive symptom scores at age 23, when the outcome is crude symptom score and logged symptom score, respectively.**

|  |  | **Outcome: symptom score** | | **Outcome: log(symptom score)** | |
| --- | --- | --- | --- | --- | --- |
| **Sex** | **Model** | **Coefficient** | **(95% CI)** | **Coefficient** | **(95% CI)** |
| **Men** | 1 (Crude) | 1.03 | (0.02, 2.03) | 0.18 | (0.04, 0.32) |
|  | 2 (Adjusted A*) | 0.21 | (-0.74, 1.17) | 0.06 | (-0.07, 0.19) |
|  | 3 (Adjusted B**) | 0.17 | (-0.80, 1.14) | 0.05 | (-0.08, 0.18) |
|  | 4 (IPTW) | 0.27 | (-0.73, 1.27) | 0.05 | (-0.16, 0.27) |
| **Women** | 1 (Crude) | 2.26 | (1.52, 3.00) | 0.31 | (0.21, 0.41) |
|  | 2 (Adjusted A*) | 1.42 | (0.74, 2.11) | 0.23 | (0.13, 0.34) |
|  | 3 (Adjusted B**) | 1.33 | (0.64, 2.01) | 0.22 | (0.12, 0.33) |
|  | 4 (IPTW) | 1.26 | (0.54, 1.98) | 0.17 | (0.01, 0.34) |

IPTW = Inverse propensity treatment weighting

Statistics are pooled from 50 multiply imputed datasets using Rubin’s rules, as described in Methods.

*Adjusted for depressive symptom score at age 16, ethnicity, socioeconomic status, and dummy variables for childhood emotional abuse, physical abuse, sexual abuse, and emotional neglect.

**As in Adjusted A*, additionally adjusted for sexual minority status, anxiety, extreme parental monitoring, anti-social behaviour, smoking, cannabis use, illicit (non-cannabis) drug use, hazardous alcohol use, bullying, witnessing domestic violence, parental mental health problem, parental substance abuse, parental criminal conviction, and parental separation.

**Table S3. Comparison of estimated coefficients from difference-in-difference analyses investigating the relationship between IPVA victimisation at age 18-21 and depressive symptom scores at ages 16 and 23, when the outcome if crude symptom score and logged symptom score, respectively.**

|  | | **Outcome: symptom score** | | | **Outcome: log(symptom score)** | | |
| --- | --- | --- | --- | --- | --- | --- | --- |
| **Sex** | **Term** | **Coefficient** | **(95% CI)** | **p-value** | **Coefficient** | **(95% CI)** | **p-value** |
| Men | Victimisation | 1.56 | (0.62, 2.50) | 0.001 | 0.29 | (0.15, 0.44) | <0.001 |
|  | Time | 1.53 | (0.95, 2.11) | <0.001 | 0.29 | (0.20, 0.38) | <0.001 |
|  | Victimisation*Time | 0.02 | (-1.29, 1.33) | 0.972 | -0.05 | (-0.25, 0.16) | 0.665 |
| Women | Victimisation | 1.61 | (0.92, 2.30) | <0.001 | 0.25 | (0.16, 0.35) | <0.001 |
|  | Time | 0.28 | (-0.22, 0.77) | 0.275 | 0.05 | (-0.02, 0.12) | 0.177 |
|  | Victimisation*Time | 0.27 | (-0.73, 1.26) | 0.598 | 0.02 | (-0.12, 0.16) | 0.758 |

CI = Confidence interval

Statistics are pooled from 50 multiply imputed datasets using Rubin’s rules, as described in Methods.

**Table S4. Percentage change in geometric mean depressive symptom scores for IPVA victimisation at age 18-21 (vs. no victimisation) from different regression models within different participant samples.**

|  | | **Main analysis sample**  **(N: Men = 1,039, Women = 1,753)** | | **Complete cases only**  **(N: Men = 719, Women = 1,329)** | | **Including those victimised by age 17**  **(N: Men = 1204, Women = 2075)** | | **Including only those who had had a romantic encounter by age 17**  **(N: Men = 894, Women = 1,528)** | |
| --- | --- | --- | --- | --- | --- | --- | --- | --- | --- |
| **Sex** | **Model** | **% change** | **(95% CI)** | **% change** | **(95% CI)** | **% change** | **(95% CI)** | **% change** | **(95% CI)** |
| **Men** | 1 (Crude) | 20 | (4, 37) | 29 | (11, 51) | 26 | (11, 42) | 12 | (-1, 27) |
|  | 2 (Adjusted A*) | 6 | (-7, 21) | 24 | (0, 54) | 9 | (-3, 23) | 13 | (-1, 28) |
|  | 3 (Adjusted B**) | 6 | (-7, 20) | 18 | (-16, 67) | 10 | (-2, 24) | 10 | (-10, 35) |
|  | 4 (IPTW) | 6 | (-15, 31) | -9 | (-33, 22) | 26 | (10, 44) | 14 | (-8, 41) |
| **Women** | 1 (Crude) | 36 | (23, 51) | 27 | (15, 41) | 42 | (29, 55) | 24 | (13, 37) |
|  | 2 (Adjusted A*) | 26 | (13, 40) | 14 | (0, 31) | 24 | (13, 35) | 22 | (11, 35) |
|  | 3 (Adjusted B**) | 25 | (12, 39) | 29 | (3, 62) | 22 | (11, 34) | 22 | (4, 42) |
|  | 4 (IPTW) | 19 | (1, 40) | 33 | (7, 65) | 43 | (29, 58) | 23 | (11, 35) |

CI = Confidence interval; IPTW = Inverse probability treatment weighting

Statistics for all samples except ‘Complete cases only’ are pooled from 50 multiply imputed datasets using Rubin’s rules, as described in Methods. % change calculated from the estimated coefficient for victimisation category in each model, as % change = [exp(coefficient)-1]*100.

*Adjusted for logged depressive symptom score at age 16, ethnicity, socioeconomic status, and dummy variables for childhood emotional abuse, physical abuse, sexual abuse, and emotional neglect.

**As in Adjusted A*, additionally adjusted for sexual minority status, anxiety, extreme parental monitoring, anti-social behaviour, smoking, cannabis use, illicit (non-cannabis) drug use, hazardous alcohol use, bullying, witnessing domestic violence, parental mental health problem, parental substance abuse, parental criminal conviction, and parental separation.

**Table S5. Estimates from difference-in-difference analysis investigating the relationship between IPVA victimisation at age 18-21 and logged depressive symptom scores at ages 16 and 23, on complete cases only.**

|  |  | **Main analysis sample**  **(N: Men = 1,039, Women = 1,753)** | | | **Complete cases only**  **(N: Men = 719, Women = 1,329)** | | | | **Including those victimised by age 17**  **(N: Men = 1204, Women = 2075)** | | | | **Including only those who had had a romantic encounter by age 17**  **(N: Men = 894, Women = 1,528)** | | |
| --- | --- | --- | --- | --- | --- | --- | --- | --- | --- | --- | --- | --- | --- | --- | --- |
| **Sex** | **Term** | **% change** | **(95% CI)** | **p-value** | | **% change** | **(95% CI)** | **p-value** | | **% change** | **(95% CI)** | **p-value** | **% change** | **(95% CI)** | **p-value** |
| **Men** | Victimisation | 34 | (16, 55) | <0.001 | | 31 | (13, 51) | <0.001 | | 36 | (19, 56) | <0.001 | 36 | (19, 56) | <0.001 |
|  | Time | 34 | (22, 47) | <0.001 | | 35 | (24, 49) | <0.001 | | 34 | (22, 46) | <0.001 | 34 | (22, 46) | <0.001 |
|  | Victimisation*Time | -5 | (-23, 18) | 0.665 | | -1 | (-20, 22) | 0.923 | | -5 | (-20, 14) | 0.594 | -5 | (-20, 14) | 0.594 |
| **Women** | Victimisation | 29 | (17, 42) | <0.001 | | 28 | (16, 42) | <0.001 | | 34 | (23, 45) | <0.001 | 33 | (22, 45) | <0.001 |
|  | Time | 5 | (-2, 13) | 0.177 | | 5 | (-2, 13) | 0.175 | | 4 | (-2, 12) | 0.199 | 4 | (-3, 12) | 0.231 |
|  | Victimisation*Time | 2 | (-11, 18) | 0.758 | | -1 | (-14, 14) | 0.909 | | 4 | (-8, 18) | 0.537 | 5 | (-7, 18) | 0.461 |

CI = Confidence interval

ϮIn geometric mean. % change calculated from the estimated coefficient for victimisation in each model, as % change = [exp(coefficient)-1]*100.

10.	Bachman JG. Youth in Transition II: The impact of family background and intelligence on tenth-grade boys. . Ann Arbor, MI: Institute of Social Research; 1970.

11.	Khambati N, Mahedy L, Heron J, Emond A. Educational and emotional health outcomes in adolescence following maltreatment in early childhood: A population-based study of protective factors. Child abuse & neglect. 2018;81:343-53.

12.	Harrison L, Wilson S, Munafo MR. Exploring the associations between sleep problems and chronic musculoskeletal pain in adolescents: a prospective cohort study. Pain Res Manag. 2014;19(5):e139-45.

13.	Lewis G, Pelosi AJ, Araya R, Dunn G. Measuring psychiatric disorder in the community: a standardized assessment for use by lay interviewers. Psychological medicine. 1992;22(2):465-86.

14.	Herbert A, Heron J, Barter C, Szilassy E, Barnes M, Howe LD, et al. Risk factors for intimate partner violence and abuse among adolescents and young adults: findings from a UK population-based cohort: Extended Data. 2020.
